# Exploring Cognitive and Neuroimaging Profiles of Dementia Subtypes of Individuals with Dementia in the Democratic Republic of Congo

**DOI:** 10.1101/2024.12.17.24319162

**Authors:** Jean Ikanga, Saranya Sundaram Patel, Megan Schwinne, Caterina Obenauf, Emmanuel Epenge, Guy Gikelekele, Nathan Tshengele, Immaculee Kavugho, Samuel Mampunza, Lelo Mananga, Charlotte E. Teunissen, Julio C. Rojas, Brandon Chan, Argentina Lario Lago, Joel H. Kramer, Adam L. Boxer, Andreas Jeromin, Emile Omba, Alvaro Alonso, Alden L. Gross

**Author notes:** Corresponding author. Please send general questions about the study to Dr. Jean Ikanga., Jean N. Ikanga, Ph.D., Department of Rehabilitation Medicine Emory University, 1441 Clifton Rd NE Atlanta, GA 30322, USA.

## Abstract

**Objective:** The 2024 Alzheimer’s Association (AA) research diagnostic criteria for Alzheimer’s Disease (AD) considers fluid biomarkers, including promising blood-based biomarkers for detecting AD. This study aims to identify dementia subtypes and their cognitive and neuroimaging profiles in older adults with dementia in the Democratic Republic of Congo (DRC) using biomarkers and clinical data.

**Methods:** Forty-five individuals with dementia over 65 years old were evaluated using the Community Screening Instrument for Dementia and the informant-based Alzheimer’s Questionnaire. Core AD biomarkers (Aβ42/40 and p-tau181) and non-specific neurodegeneration biomarkers (NfL, GFAP) were measured in blood plasma. Neuroimaging structures were assessed using magnetic resonance imaging (MRI). Dementia subtypes were determined based on plasma biomarker pathology and vascular markers. Biomarker cutoff scores were identified to optimize sensitivity and specificity. Individuals were stratified into one of four dementia subtypes – AD only, non-AD vascular, non-AD other, or mixed – based on combinations of abnormalities in these markers.

**Results:** Among the 45 individuals with dementia, mixed dementia had the highest prevalence (42.4%), followed by AD-only (24.4%), non-AD other dementia (22.2%), and non-AD vascular dementia subtypes (11.1%). Both cognitive and neuroimaging profiles aligned poorly with biomarker classifications in the full sample. Cognitive tests varied across dementia subtypes. The cognitive profile of the AD-only and mixed groups suggested relatively low cognitive performance, while the non-AD and other groups had the best scores on average.

**Conclusion:** Consistent with studies in other settings, our preliminary findings suggest that neurodegenerative plasma biomarkers may help to identify dementia subtypes and provide insight into cognitive and neuroimaging profiles among older adults in the DRC.

## INTRODUCTION

Alzheimer’s disease (AD) is the most common neurodegenerative disease, with pathology characterized by the accumulation of amyloid-beta (Aβ) plaques and neurofibrillary tangles composed of hyperphosphorylated tau protein.^1^ With the advancements in assay technology, plasma biomarkers have increasingly been shown to have potential for the detection and monitorization of AD, increasing accessibility beyond catchment areas of major medical centers.^2,3^ Current revised 2024 Alzheimer’s Association (AA) criteria distinguish three broad categories of AD fluid biomarkers related to AD pathogenesis: (1) core AD fluid biomarkers (the CSF ratio of amyloid-β [Aβ42/40], phosphorylated and secreted AD tau (p-tau 217, p-tau-181, and p-tau 231)), (2) non-specific biomarkers involved in other neurodegenerative pathology, including neurofilament light (NfL) and glial fibrillary acidic protein (GFAP), and (3) biomarkers of non-AD pathology (vascular brain injury, alpha-synuclein [αSyn]).^4^ Identifying plasma biomarkers for underlying pathologies of dementia can especially benefit prodromal or pre-clinical stages, for which current and emerging disease-modifying therapies are more likely to be effective.^5^

Using blood biomarkers known to provide early indication of a disease may facilitate more timely diagnosis for patients exhibiting early symptoms, particularly in early-onset and atypical presentations. Blood-based biomarkers in AD are associated with both early indicators of cognitive decline and longitudinal cognitive outcomes.^2^ For example, lower plasma Aβ42/Aβ40 ratios correlate with higher amyloid plaque burden and cognitive impairment and can be detected in preclinical disease stages,^6^ making it useful for early diagnosis and tracking disease progression.^7^ Elevated levels of p-tau181 and p-tau217 are observed in AD, serving as indicators of both early and late stages of AD.^8,9^ Plasma p-tau increases in early symptomatic stages, aligning with clinical transition from mild cognitive impairment (MCI) to AD dementia.^10^ NfL is a marker of axonal damage; while less specific, elevated levels of NfL reflect more widespread neuronal damage.^11^ GFAP reflects astrocytic activation and neuroinflammation, with increased levels observed in AD. GFAP may be used to complement other biomarkers to enhance diagnostic accuracy, particularly in advanced stages.^12^ However, additional data are still needed to demonstrate the utility and validity of blood biomarkers in diverse clinical cohorts and to accurately detect disease profiles, particularly given overlapping symptom profiles across different pathologies. For example, vascular damage and protein alterations are present in most forms of dementia, which adds a layer of uncertainty to diagnosis given the potential for mixed dementia pathology.^13^ In vascular disease, NfL may also be elevated as axonal injury can be seen in cerebrovascular disease. NfL concentrations reflect acute and chronic cerebrovascular injury, which is useful for both early detection and monitoring progression.^14^ In addition to plasma biomarkers, structural neuroimaging may also provide important additional diagnostic data. Specifically, the entorhinal cortex and hippocampal regions are particularly affected in the early stages of AD.^1,15^ Hippocampal volume loss is a feature differentiating AD dementia from other dementias, such frontotemporal dementia (FTD) and vascular dementia (VaD), and is closely linked to the course of AD.^16–17^

A significant caveat is that most research involving neurodegenerative biomarkers in AD primarily have been conducted using Western cohorts. Studies have shown that CSF biomarkers, such as reduced levels of Aβ42 and p-tau, correlate with AD pathology and can aid in distinguishing AD from other forms of dementia,^18^ but less is known about the biomarker and neuroimaging parameters and profiles in diverse populations, particularly in Sub-Saharan African (SSA) populations.

The current study aims to explore dementia subtypes based on blood-based biomarkers and vascular factors, and their neuroimaging and cognitive profiles in adult individuals with clinical dementia in Kinshasa, Democratic Republic of Congo (DRC), in SSA. We expected that there is higher prevalence of participants with non-AD pathologies compared to those with AD dementia subtype. Based on previous studies that have linked amyloid-β deposition, tau protein, and neurodegeneration (NFl) accumulations with impairments in language, learning and memory, and executive function, we hypothesized that the cognitive patterns aligning with neurodegenerative biomarkers are characterized by deficits in these cognitive domains ^6–12,19^. Similarly, since amyloid-β deposition and tau protein accumulation in the brain are associated with atrophy in the hippocampus, temporal lobe, medial temporal, and entorhinal cortex, we expected that the neuroimaging patterns that align with neurodegenerative biomarkers are characterized by atrophy in these structures ^1,15–17^. A general comparison of cognitive deficits and brain atrophy reveals more severe and distinct patterns of deficits and atrophy in Alzheimer’s disease (AD) participants, followed by those with mixed dementia, and vascular dementia.

## METHODS

### Study population

Participants of this study are community-dwellers from Kinshasa/DRC diagnosed with dementia and selected from a prevalence study of dementia.^20^ Study design details have been published previously.^20^ Briefly, participants were included if they were at least 65 years or older, had a family member or close friend to serve as an informant, and fluent in French or Lingala. We excluded individuals who had history of schizophrenia, neurological, or other medical conditions potentially affecting the central nervous system (CNS), yielding a sample of 1,432 eligible participants. To establish neurological status in the absence of established diagnostic criteria for AD in Sub-Saharan Africa (SSA), we screened eligible participants using the Alzheimer’s Questionnaire (AQ)^21^ and the Community Screening Instrument for Dementia (CSID).^22–23^ The AQ assesses activities of daily living and symptoms of AD in participants.^21^ The CSID Questionnaire, used in several SSA dementia studies,^24–26^ was used to screen cognitive abilities.

Based on cognitive and functional deficits per the Diagnostic and Statistical Manual of Mental Disorders, Fifth Edition, Text Revision (DSM-5-TR) diagnostic criteria,^27^ we classified eligible participants using CSID cut-offs from a previous study conducted in Congo-Brazzaville, the closest city from Kinshasa.^28^ Similar to our prior study,^20^ eligible participants were classified using CSID and AQ scores (see Figure 1) which resulted in 1,161 individuals being excluded based on their having only mild neurocognitive disorder (MND) or subjective cognitive impairment.

**Figure 1.**
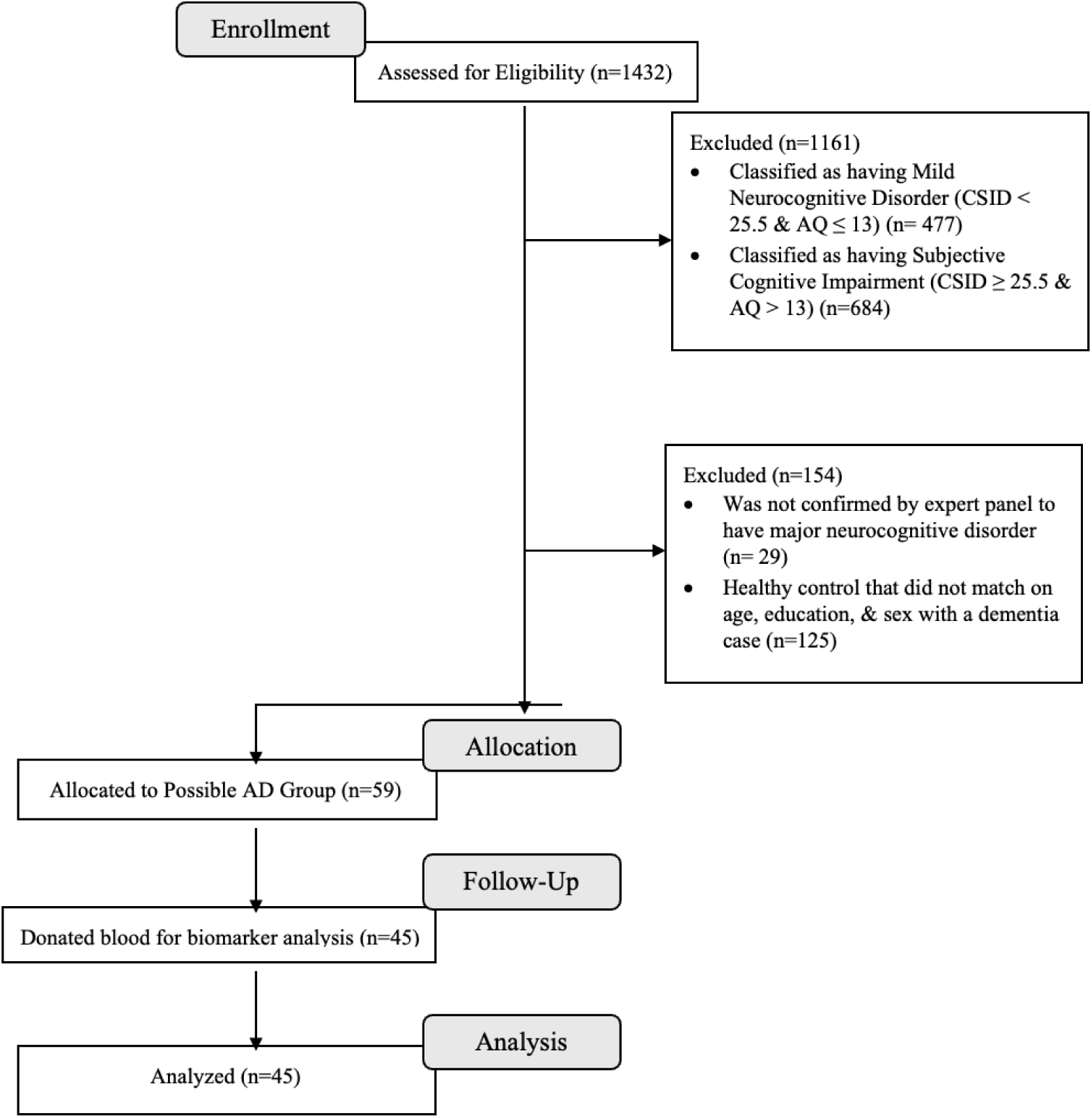
Flow Chart of Recruitment Status from those assessed for eligibility at enrollment (n=1432) to the individuals that were allocated to the dementia and donated blood for biomarkers (n=45)

A panel consisting of a neurologist (EE), psychiatrist (GG) and neuropsychologist (JI) reviewed screening tests, clinical interview, and neurological examination of 271 subjects, of whom 59 from 88 were confirmed with a diagnosis of dementia and 58 from 183 were considered HC. Of these 117 participants, 29 refused to provide blood samples, leaving 85 participants (75%) in whom plasma biomarkers were obtained (45 dementia and 40 HC) who were matched on age, education, and sex. For the present analysis, only participants with dementia were included (See Figure 1). Written informed consent was obtained prior to participants’ undergoing any study procedures. Participants were financially compensated for their time. The procedures were approved by the Ethics Committee/Institutional Review Boards of the University of Kinshasa and Emory University.

### Procedure

Participants underwent a comprehensive clinical evaluation, including cognitive testing, self-report questionnaires, and standard psychiatric and neurological evaluations. Subjects were interviewed to obtain demographic, socioeconomic, and medical history and were subsequently administered cognitive testing with African Neuropsychological Battery (ANB) subtests.

### Measures

#### Plasma biomarkers

Blood samples were drawn at the Medical Center of Kinshasa (CMK) blood laboratory by antecubital venipuncture into dipotassium ethylene diamine tetra acetic acid (K_2_ EDTA) tubes. Samples were centrifuged within 15 minutes at 1800 g house temperature, and 5 mL of plasma was aliquoted into 0.5 mL polypropylene tubes and stored initially at -20° C for less than a week and stored in a -80 °C freezer for longer term storage at a CMK laboratory. These aliquots were shipped frozen on dry ice to Emory University for storage and then to University of California San Francisco (UCSF) for measurements.

Plasma biomarker concentrations were measured using commercially available Neurology 4-PLEX E (Aβ40, Aβ42, NfL, and GFAP; lot #503819), P-Tau181 (P-Tau181 v2; lot #503732), IL-1b (lot #503806) and IL-10 (IL-10 2.0, lot #503533) Quanterix kits on the Simoa HD-X platform (Billerica, MA) at UCSF. P-tau217 was measured using the proprietary ALZpath pTau-217 CARe Advantage kit (lot #MAB231122, ALZpath, Inc.) on the Simoa HD-X platform. The instrument operator was blinded to clinical variables. All analytes were measured in duplicate, except for IL-1b, which was measured as a singlicate due to low sample availability. ^45–47^ For Aβ40, Aβ42, NfL, and GFAP, all samples were measured above the lower limit of quantification (LLOQ) of 1.02 pg/mL, 0.378 pg/mL, 0.4 pg/mL and 2.89 pg/mL, respectively. The average coefficient of variation (CV) for Aβ40, Aβ42, NfL, and GFAP were 6.0%, 6.5%, 5% and 4.6%, respectively. For IL-1b and IL-10, the LLOQ were 0.083 pg/mL and 0.021 pg/mL, respectively. The average CV for IL-10 was 6.1%. For P-tau217 the LLOQ was 0.024 pg/mL and the average CV was 19.8%.^45^

#### Neuroimaging

All subjects were imaged on a 1.5 Tesla MRI unit (Siemens, Magneton Sonata) scanner at HJ Hospitals in Kinshasa using the same standardized imaging acquisition protocol based on the Alzheimer’s Disease Research Center (ADRC) protocol of Emory University.^30^ This consisted of sagittal volumetric T1-weighted (MPRAGE), coronal T2-weighted, and axial diffusion-weighted, T2-weighted, and T2-FLAIR sequences. Typical acquisition parameters for the MPRAGE sequence were TR = 2200 ms, minimum full TE, TI = 1000 ms, flip angle = 8°, FOV = 25 cm, with a 192 × 184 acquisition matrix, yielding a voxel size of approximately 1.25 × 1.25 × 1.2 mm.

Images were reviewed by a subspecialty certified neuroradiologist (AMS) with 14 years of experience. White matter hyperintensities were graded according to the Age-Related White Matter Changes (ARWMC) scale.^31^ The number of chronic brain parenchymal microhemorrhages were recorded. MPRAGE images were reoriented into the oblique coronal plane orthogonal to the principal axis of the hippocampal formation, and medial temporal lobe atrophy (MTLA)^32^ and entorhinal cortex atrophy (EriCa)^33^ scores were assessed. Finally, the presence or absence of any additional abnormalities was noted, and patients were excluded if neuroimaging evidence indicated an etiology other than probable AD (e.g., presence of a brain tumor).

#### Quantitative volumetric analysis using Freesurfer

The 3D T1 images were segmented using Freesurfer (v.6, MGH, MA), which includes a full processing stream for MR imaging data that involves skull-stripping, bias field correction, registration, and anatomical segmentation as well as cortical surface reconstruction, registration, and parcellation. Regional brain volume for both cortical and subcortical brain regions were calculated. The left and right hippocampal volume were averaged. Interindividual variation in head size were accounted for in further statistical analysis by controlling for the effects of the total intracranial volume.

### Determination of Dementia Subtypes

Dementia subtypes were determined based on the plasma biomarkers (Aβ_42/40_, p-tau_181_, NfL, GFAP), alongside vascular markers, hemoglobin A1c (HbA1c), blood pressure, and total cholesterol. Given lack of established AD biomarker thresholds in the DRC/SSA, determination of biomarker thresholds was informed by prior analysis conducted by Ikanga and colleagues (2024)^29^ in which areas under the curve were calculated to predict diagnostic accuracy of biomarkers on neurological status (healthy or suspected AD).^30^ Thresholds for vascular markers, HbA1c, hypertension, and hypercholesterolemia, were sourced from existing literature.^34–37^ Individuals with elevated HbA1c (≥6.5%), blood pressure (systolic ≥130 mmHg or diastolic ≥80 mmHg), or total cholesterol (≥200 mg/dL) were deemed to have dementia of potential vascular etiology. Subsequently, individuals were classified into one of four dementia subtypes – AD only, non-AD vascular, non-AD other, or mixture – based on their presence or absence of these biomarkers (Table 1).

**Table 1.** Organization of Biomarkers into Pathological Subtypes of Dementia Utilizing Ikanga and al (2024)^29^ Threshold for Core AD Biomarkers.

| Biomarker | Threshold | Pathological Type |  |  |  |
| --- | --- | --- | --- | --- | --- |
|  |  | AD only | Non-AD Vascular | Non-AD Other | Mixed |
| Core AD Biomarkers |  | Present | Absent | Absent | Present |
| Decreased A$\beta_{42/40}$ | $\leq 0.061$ pg/ml | | | | |
| Increased p-tau$_{181}$ | $\geq 4.50$ pg/ml | | | | |
| Non-specific AD biomarker |  | Optional | Present | Optional | Optional |
| Increased GFAP | $\geq 176.0$ pg/ml | | | | |
| Vascular Markers |  | Absent | Present | Optional | Present |
| High HbA1c | $\geq 6.5\%$ | | | | |
| Hypertension | SBP $\geq 130$ or DBP $\geq 80$ mmHg | | | | |
| Hypercholesterolemia | TC $\geq 200$ mg/dL | | | | |

\*Note-in merged rows, if a biomarker is required to be present, only one, but not limited to one biomarker needs to be present. However, all biomarkers required to be absent must be absent; for example, AD only subtype requires either Decreased A $\beta_{42/40}$ , Increased p-tau<sub>181</sub>, or both. Increased GFAP is optional, but all vascular markers must be absent.
Abbreviations: AD = Alzheimer's Disease, A $\beta_{42/40}$ = ratio of amyloid beta 42 and amyloid beta 40, p-tau<sub>181</sub> = phosphorylated tau protein 181, NfL = neurofilament light chain, GFAP = glial fibrillary acidic protein, HbA1c = hemoglobin A1c, SBP/DBP = systolic/diastolic blood pressure, TC = total cholesterol

### Statistical Analyses

Descriptive statistics were employed to summarize the data, with continuous variables reported as means and standard deviations, and categorical variables reported as frequencies and row percentages. We used linear regression models to compare differences in demographics, biomarkers, vascular markers, neuroimaging measures, and cognitive tests by dementia subtype. Models were adjusted for age, gender, years of education, total intracranial volume (for neuroimaging variables), and Geriatric Depression Scale (GDS) score. Subsequently, Dunn’s post hoc test for pairwise comparisons was conducted to explore differences in neuroimaging and cognitive assessment measure between biomarker-defined dementia subtypes. Results were evaluated with a significance set at *p*<.05. Statistical analysis was conducted using R version 4 statistical software.

## RESULTS

Demographic data, neurodegenerative plasma biomarkers, vascular markers, neuroimaging, and cognitive characteristics are presented in Table 2. The sample comprised 45 clinically adjudicated dementia participants, of whom 20 (44%) were males, with an average age of 73.8 years (SD = 8 years) and an average of 7.4 years of education (SD = 5 years). Clinically, the sample exhibited high symptoms of depression (GDS = 7.5), and 58% of the participants had clinical hypertension (see Table 2).

**Table 2.** Characteristics of the Study Sample.

| Characteristic, $\mu$ ($\sigma$) | **Clinical**<br>**Dementia**<br>(n=45) |
| --- | --- |
| Age (years) | 73.8 (8) |
| Male (n, %) | 20 (44%) |
| Education (years) | 7.4 (5) |
| GDS Score | 7.5 (3.5) |
| **Biomarkers** |  |
| A$\beta_{42/40}$ | 0.06 (0.03) |
| p-tau 181 (pg/ml) | 3.0 (2) |
| NfL (pg/ml) | 62.7 (41) |
| GFAP (pg/ml) | 241.0 (144) |
| **Vascular Markers** |  |
| HbA1c (%) | 5.6 (0.7) |
| Hypertension (n, %) | 26 (58%) |
| High Cholesterol (n, %) | 1 (2%) |
| **Neuroimaging Measures** |  |
| Intracranial Volume (mm<sup>3</sup>) | 1433637<br>(277941) |
| Left Hippocampal Volume (mm<sup>3</sup>) | 2970 (535) |
| Right Hippocampal Volume (mm<sup>3</sup>) | 2973 (573) |
| Left Entorhinal Cortex Volume (mm<sup>3</sup>) | 1525 (573) |
| Right Entorhinal Cortex Volume (mm<sup>3</sup>) | 1642 (568) |
| White Matter Hyperintensity | 70.0 (2.6) |
| Microhemorrhage | 0.69 (1.5) |
| Mesial Temporal Atrophy Score | 2.3 (1.1) |
| Entorhinal Cortex Atrophy Score | 1.7 (0.78) |
| **Cognitive Tests** |  |
| CSID | 19.6 (5.6) |
| AQ | 19.3 (4.0) |
| African Naming Test | 15.5 (7.2) |
| ALMT Trial 1 | 2.6 (1.7) |
| AVMT Trial 1 | 1.3 (1.6) |
| ALMT Trial 3 | 4.0 (1.9) |
| AVMT Trial 3 | 1.9 (1.9) |
| ALMT Recall | 0.31 (0.6) |
| AVMT Recall | 1.0 (1.7) |
| Proverb Test | 2.5 (2.2) |
| Card Game Wins | 21.7 (7.0) |

\*GDS = Geriatric Depression Score; NfL = Neurofilament Light; GFAP = Glial Fibrillary Acidic Protein; CSID = Community Screening Instrument for Dementia; AQ = Alzheimer's Questionnaire; ALMT = African List Memory Test; AVMT = African Visuospatial Memory Test.

Table 3 presents the dementia subtype defined by neurodegenerative plasma biomarkers using Ikanga and colleagues’ threshold^29^. As anticipated, there is a higher prevalence of mixed dementia, followed by AD-only, non-AD other dementia, and non-AD vascular dementia patterns.

**Table 3.**
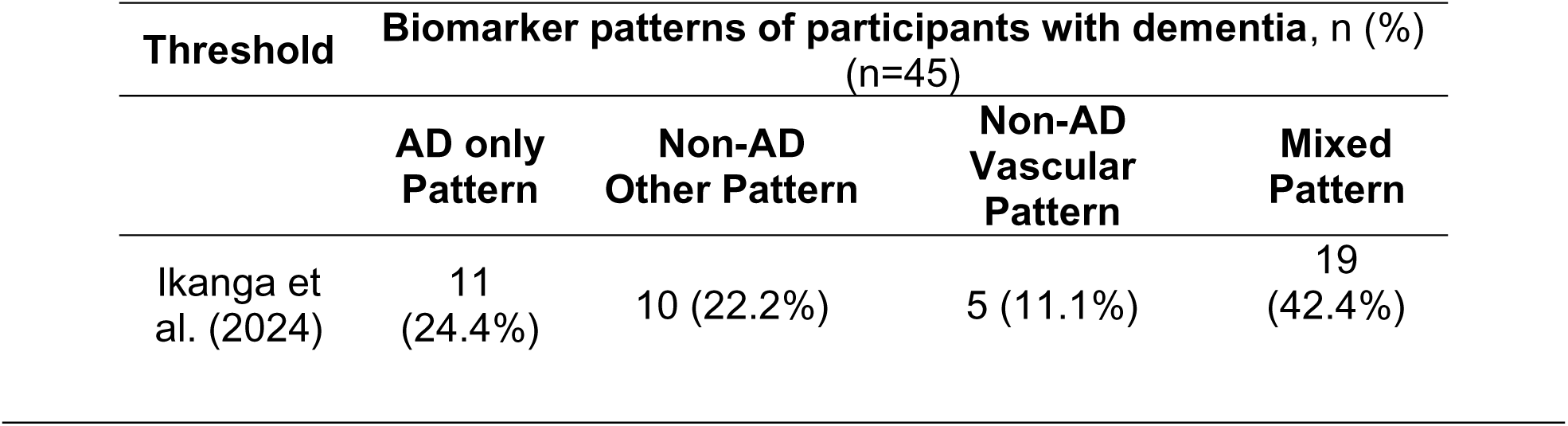
Clinical Dementia Subtype based on biomarker patterns.

Table 4 presents the cognitive profiles for each dementia subtype based on the cutoff criteria established by Ikanga and colleagues (2024)^29^. We excluded the vascular dementia subtype from these analyses due to the small sample size (only 5 participants). Cognitively, there were no clinically or statistically significant differences between the dementia subtypes. The cognitive profiles do not align well with biomarker-based dementia subtypes.

**Table 4.** Cognitive Profile for Dementia Subtypes.

| **Cognitive Test** | **Dementia Subtype, $\mu$ ($\sigma$)** |  |  | **p-value** |
| --- | --- | --- | --- | --- |
|  | **AD Only**<br>**(n=11)** | **Non-AD**<br>**Other**<br>**(n=10)** | **Mixture**<br>**(n=19)** |  |
| African Naming Test | 12.6 (6.44) | 19.5 (5.85) | 14.9 (7.67) | 0.099 |
| ALMT Trial 1 | 2.22 (0.97) | 3.30 (1.34) | 2.89 (1.91) | 0.32 |
| AVMT Trial 1 | 1.11 (1.27) | 1.90 (1.66) | 1.39 (1.82) | 0.50 |
| ALMT Trial 3 | 3.22 (1.39) | 4.90 (1.66) | 4.22 (2.02) | 0.16 |
| AVMT Trial 3 | 1.11 (1.05) | 2.60 (2.50) | 2.11 (1.84) | 0.11 |
| ALMT Recall | 0.00 (0.00) | 0.30 (0.67) | 0.50 (0.79) | 0.22 |
| AVMT Recall | 0.22 (0.44) | 1.70 (2.41) | 1.11 (1.60) | 0.15 |
| Proverb Test | 2.00 (1.22) | 4.00 (3.37) | 2.11 (1.64) | 0.056 |
| Card Game Wins | 23.8 (8.17) | 21.9 (6.88) | 16.4 (4.93) | 0.76 |

Table 5 presents the neuroimaging profile for each dementia subtype based on Ikanga and colleagues’ threshold^29^. As in the previous analyses, we did not include the vascular dementia subtype because there are only 5 participants in this subtype. The neuroimaging profiles do not align well with biomarker-based dementia subtypes. The AD group showed reduced scores in many neuroanatomical structures compared to other dementia subtypes. There was a statistical difference in left hippocampal volume between various dementia subtypes, mostly between AD-only and non-AD other subtypes, and between AD-only and mixed subtypes. There was a trend in terms of microhemorrhage between dementia subtypes (see Table 5).

**Table 5:** Neuroimaging Profile for Dementia Subtypes.

| **Neurological Measure** | **Dementia Subtype, mean (SD)** |  |  | **p-value** |
| --- | --- | --- | --- | --- |
|  | **AD Only**<br>**(n=11)** | **Non-AD**<br>**Other** | **Mixture**<br>**(n=19)** |  |

|  | (n=10) |  |  |  |
| --- | --- | --- | --- | --- |
| Intracranial Volume | 1595574<br>(506956) | 1419302<br>(162980) | 1364517<br>(139986) | 0.11 |
| Left Hippocampus*† | 2523<br>(355) | 3122<br>(408) | 3142<br>(611) | 0.006 |
| Right Hippocampus | 2596<br>(210) | 3011<br>(564) | 3122<br>(730) | 0.13 |
| Left Entorhinal Cortex | 1248<br>(431) | 1696<br>(504) | 1596<br>(620) | 0.12 |
| Right Entorhinal Cortex | 1433<br>(504) | 1732<br>(443) | 1750<br>(707) | 0.24 |
| White Matter Hyperintensity | 70.3<br>(2.97) | 71.0<br>(2.39) | 69.3<br>(2.62) | 0.36 |
| Microhemorrhage | 0 (0) | 5 (50%) | 3 (16%) | 0.053 |
| Mesial Temporal Atrophy<br>Score | 2.78<br>(0.83) | 2.10 (0.99) | 1.77<br>(1.01) | 0.081 |
| Entorhinal Cortex Atrophy<br>Score | 1.78<br>(0.83) | 1.60 (0.70) | 1.54<br>(0.88) | 0.66 |

\* Statistically significant difference between AD-only and non-AD other subtypes
† Statistically significant difference between AD-only and mixture subtypes
Note: Presence of microhemorrhages is a dichotomous variable, represented as n (%)

## DISCUSSION

This study primarily aimed to evaluate the feasibility of using neurodegenerative plasma biomarkers to characterize dementia subtypes and describe their cognitive and neuroimaging profiles in a novel sample of older adults with clinical dementia in SSA. Experts have reported gaps in neuropsychological testing instruments, diagnostic procedures, fluid biomarkers, and neuropathological correlative studies.^38^ This exploratory study aimed to address the gap in plasma biomarkers in the DRC/SSA.

Despite the absence of a gold standard threshold for neurodegenerative fluid biomarkers in the DRC/SSA, we investigated the cognitive and neuroimaging profiles of adults with dementia in the DRC/SSA, using plasma neurodegenerative biomarkers. We found a high prevalence of mixed, followed by AD only, non-AD other dementia, and non-AD vascular dementia patterns, despite cultural, racial, and geographic differences. These results contrast with Western findings, which indicate that the most prevalent dementias among older adults (65 years and over) are Alzheimer’s Disease (60-80% of cases), vascular dementia (10-20% of cases), mixed dementia (5-15% of cases), and other dementias, such as dementia with Lewy bodies (2-5% of cases), dementia associated with Parkinson’s disease (3.6% of cases), and frontotemporal dementia (2-5% of cases).^39–41^

These differences in classifying dementia based on clinical and biological markers can be explained by the heterogeneous and continuous nature of Alzheimer’s disease, which is complex to characterize.^42^ Clinical adjudication relies on medical history, neuropsychological assessments, cognitive symptoms, and behavioral changes, which can be subjective and prone to variability among clinicians.^43^ Fluid biomarkers can assess specific proteins or molecules, detect biological changes, and provide objective, quantitative measures to refine clinical diagnosis. Therefore, fluid biomarkers can identify pre-symptomatic or prodromal stages of dementia.^44^ Thus, there could be changes in the classification of AD prevalence.

Contrary to our second hypothesis, which predicted that the neuroimaging profile would align better with biomarker-based dementia subtypes than with the cognitive profile, we found that neither cognitive nor neuroimaging profile tracked well with plasma biomarkers. Our results showed that cognitive tests did not track well with biomarker classifications among those clinical dementia participants. The cognitive profile in the AD-only and Mixed groups suggests relatively low cognitive performance, while the Non-AD Other group has some of the best scores on average. Biological underpinnings may explain some further variance in cognitive profiles, particularly in the AD only group.

The neuroimaging profile appeared to track poorly with biomarker classifications among those with clinical dementia. The Mixed group seems to have a relatively preserved neuroimaging profile. Among those with clinically adjudicated dementia, the AD-only biomarker group has significantly lower volumes for some variables. This suggests that biological underpinnings might explain some further variance in the neuroimaging profile among people with dementia. In sum, blood-based biomarkers appear to show differences in cognitive and neuroimaging parameters, largely among people with AD dementia.

As noted, this is the first study to explore biomarker-based dementia subtypes and to examine cognitive and neuroimaging profiles in the DRC/SSA using culturally appropriate neuropsychological tests, neuroimaging tools, and fluid biomarkers. The exploratory findings of this study provide evidence of the usefulness of ANB tests and their importance in the algorithm for clinical adjudication of different subtypes of dementia. Our analyses also showed the importance of MRI and plasma biomarkers as diagnostic tools for dementia in SSA/DRC. Overall, the strengths of the current study include the use of culturally validated neuropsychological tests, the ability to collect neuroimaging data in participants who are not familiar with MRI, and plasma biomarkers in a population where there is resistance to donating blood for research due to fear of witchcraft. This study used a case-control design to obtain cross-sectional results.

Some limitations of this exploratory study include the modest sample size in this first DRC effort, given the cost of collection, shipping, and the analyses of plasma biomarkers, MRI scans, and the novel nature of their introduction in the DRC, which created some hesitancy for many potential participants to enroll in the study. Overall, we were pleasantly surprised by the success of our project, and we hope to recruit even larger samples in the future, and to analyze other neurodegenerative fluid biomarkers and the staging of various dementia subtypes. We are very hopeful that our work will contribute to improving clinical and biological adjudication of the accuracy of the diagnosis of AD and other neurodegenerative dementias in SSA, which will, in turn, decrease the potential diagnostic heterogeneity that might currently exist. Additionally, we only focused on participants with dementia without including other intermediary cognitive decline (e.g., MCI cohort), which could be seen as a limitation as well. The decision to exclude MCI was based on both funding limitations and the opportunity to investigate patterns of dementia that are well characterized in the Western world, as an opportunity to establish the validity of these techniques in SSA. With this success, we ultimately want to build a cohort of more diverse Congolese older adults to investigate many other fluid biomarker hypotheses tested in the West.

In conclusion, despite some limitations, the current study provides the first and preliminary patterns of dementia based on the biological definition of dementia and their cognitive and neuroimaging profiles in elderly adults with clinical dementia in Kinshasa/DRC. Future research should build on the methods and findings provided by our exploratory study to establish gold standard thresholds for different fluid biomarkers, the classification of various dementia subtypes based on these biomarkers, and the harmonization with clinical classification in probable AD and related dementia patients in SSA and DRC. Future research should also include cohorts of patients with intermediary status of cognitive decline, amnestic and non-amnestic dementia.

## Data Availability

All data produced in the present study are available upon reasonable request to the authors

## DATA AVAILABILITY STATEMENT

The raw data supporting the conclusions of this article will be made available by the authors upon appropriate request.

## ETHICS STATEMENT

The studies involving humans were approved by University of Kinshasa and Emory University. The studies were conducted in accordance with the local legislation and institutional requirements. The participants provided their written informed consent to participate in this study

## AUTHOR CONTRIBUTIONS

JI: Conceptualization, Data curation, Formal analysis, Funding acquisition, Investigation, Methodology, Project administration, Resources, Software, Supervision, Validation, Visualization, Writing – original draft, Writing – review & editing. SP: Writing – original draft, Writing – review & editing. MS: Writing – original draft, Writing – review & editing. CA: Writing – review & editing. EE: Writing – review & editing. GG: Writing – review & editing. NT: Writing – review & editing. IK: Writing – review & editing. SM: Writing – review & editing. LM: Writing – review & editing. CT: Writing – review & editing. AS: Writing – review & editing. JR: Writing – review & editing. BC: Writing – review & editing. AL: Writing – review & editing. JK: Writing – review & editing. AB: Writing – review & editing. AJ: Writing – review & editing. AG: Writing – review & editing. AA: Writing – review & editing.

## REFERENCES

1. Chavan AB, Patil SR, Patel AM, Chaugule SV, Gharal SK. A comprehensive review on Alzheimer’s disease its pathogenesis, epidemiology, diagnostics and treatment. J Res Appl Sci Biotechnol. 2023;2(4):66–72. doi:10.55544/jrasb.2.4.8

2. Assfaw AD, Schindler SE, Morris JC. Advances in blood biomarkers for Alzheimer disease (AD): A review. Kaohsiung J Med Sci. 2024;40(8):692–8. doi:10.1002/kjm2.12870

3. Palmqvist S, Tideman P, Mattsson-Carlgren N, et al. Blood biomarkers to detect Alzheimer disease in primary care and secondary care. JAMA. 2024;332(15):1245–57. doi:10.1001/jama.2024.13855

4. Jack Jr CR, Andrews SJ, Beach TG, et al. Revised criteria for the diagnosis and staging of Alzheimer’s disease. Nat Med. 2024;30(8):2121–4. doi:10.1038/s41591-024-02988-7

5. Ashton NJ, Hye A, Rajkumar AP, et al. An update on blood-based biomarkers for non-Alzheimer neurodegenerative disorders. Nat Rev Neurol. 2020;16(5):265–84. doi:10.1038/s41582-020-0348-0

6. Nakamura A, Kaneko N, Villemagne VL, et al. High performance plasma amyloid-β biomarkers for Alzheimer’s disease. Nature. 2018;554(7691):249–54. doi:10.1038/nature25456

7. Palmqvist S, Janelidze S, Stomrud E, et al. Performance of fully automated plasma assays as screening tests for Alzheimer disease–related β-amyloid status. JAMA Neurol. 2019 Sep 1;76(9):1060–9. doi:10.1001/jamaneurol.2019.1632

8. Karikari TK, Pascoal TA, Ashton NJ, et al. Blood phosphorylated tau 181 as a biomarker for Alzheimer’s disease: a diagnostic performance and prediction modelling study using data from four prospective cohorts. Lancet Neurol. 2020;19(5):422–33. doi:10.1016/S1474-4422(20)30071-5

9. Janelidze S, Stomrud E, Smith R, et al. Cerebrospinal fluid p-tau217 performs better than p-tau181 as a biomarker of Alzheimer’s disease. Nat Commun. 2020;11(1):1683. doi:10.1038/s41467-020-15436-0

10. Thijssen EH, La Joie R, Wolf A, et al. Diagnostic value of plasma phosphorylated tau181 in Alzheimer’s disease and frontotemporal lobar degeneration. Nat Medicine. 2020;26(3):387–97. doi:10.1038/s41591-020-0762-2

11. Mattsson N, Andreasson U, Zetterberg H, Blennow K, Alzheimer’s Disease Neuroimaging Initiative. Association of plasma neurofilament light with neurodegeneration in patients with Alzheimer disease. JAMA Neurol. 2017;74(5):557–66. doi:10.1001/jamaneurol.2016.6117

12. Oeckl P, Anderl-Straub S, Von Arnim CA, et al. Serum GFAP differentiates Alzheimer’s disease from frontotemporal dementia and predicts MCI-to-dementia conversion. J Neurol Neurosurg Psychiatry. 2022;93(6):659–67. doi:10.1136/jnnp-2021-328547

13. Dodge HH, Zhu J, Woltjer R, et al. Risk of incident clinical diagnosis of Alzheimer’s disease–type dementia attributable to pathology-confirmed vascular disease. Alzheimers Dement. 2017;13(6):613–23. doi:10.1016/j.jalz.2016.11.003

14. Mattsson N, Cullen NC, Andreasson U, Zetterberg H, Blennow K. Association between longitudinal plasma neurofilament light and neurodegeneration in patients with Alzheimer disease. JAMA Neurol. 2019;76(7):791–9. doi:10.1001/jamaneurol.2019.0765

15. Igarashi KM. Entorhinal cortex dysfunction in Alzheimer’s disease. Trends Neurosci. 2023;46(2):124–36. doi:10.1016/j.tins.2022.11.006

16. Maclin JM, Wang T, Xiao S. Biomarkers for the diagnosis of Alzheimer’s disease, dementia Lewy body, frontotemporal dementia and vascular dementia. Gen Psychiatr. 2019;32(1). doi:10.1136/gpsych-2019-100054

17. Killiany RJ, Hyman BT, Gomez-Isla TM, et al (2002). MRI measures of entorhinal cortex vs hippocampus in preclinical AD. Neurology 58, 1188–1196. doi: 10.1212/wnl.58.8.1188

18. van Harten AC, Wiste HJ, Weigand SD, et al. Detection of Alzheimer’s disease amyloid beta 1-42, p-tau, and t-tau assays. Alzheimers Dement. 2022 Apr;18(4):635–44. doi:10.1002/alz.12406

19. Slot RE, Sikkes SA, Berkhof J, et al. Subjective cognitive decline and rates of incident Alzheimer’s disease and non–Alzheimer’s disease dementia. Alzheimers Dement. 2019;15(3):465–76. doi:10.1016/j.jalz.2018.10.003

20. Ikanga J, Reyes A, Kaba D, et al. Prevalence of suspected dementia in a sample of adults living in Kinshasa-Democratic Republic of the Congo. Alzheimers Dement. 2023;19(9):3783–93. doi:10.1002/alz.13003

21. Malek-Ahmadi M, Davis K, Belden C, et al. Validation and diagnostic accuracy of the Alzheimer’s questionnaire. Age Ageing. 2012 May 1;41(3):396–9. doi:10.1093/ageing/afs008

22. Hall KS, Gao S, Emsley CL, Ogunniyi AO, Morgan O, Hendrie HC. Community screening interview for dementia (CSI ‘D’); performance in five disparate study sites. Int J Geriatr Psychiatry. 2000;15(6):521–31. doi: 10.1002/1099-1166(200006)15:6<521::aid-gps182>3.0.co;2-f

23. Imarhiagbe F, Ogunrin O, Ogunniyi A. Cognitive performance of hypertensive elderly Nigerians: a case control study. Afr J Med Med Sci. 2005;34(3):269–73.

24. Farombi TH, Owolabi MO, Ogunniyi A. Falls and Their Associated Risks in Parkinson’s Disease Patients in Nigeria. J Mov Disord. 2016;9(3):160–165. doi:10.14802/jmd.16011

25. Ogbimi EM, Akemokwe FM, Ogunrin O. Frequency, pattern and predictors of cognitive impairments in patients with Parkinson’s disease using the Community Screening Instrument for Dementia. Front Hum Neurosci. 2023;17:1126526. doi:10.3389/fnhum.2023.1126526

26. Akinyemi RO, Allan L, Owolabi MO, et al. Profile and determinants of vascular cognitive impairment in African stroke survivors: the CogFAST Nigeria Study. J Neurol Sci. 2014;346(1-2):241–249. doi:10.1016/j.jns.2014.08.042

27. American Psychiatric Association. Diagnostic and Statistical Manual of Mental Disorders. 5th ed., text rev. Arlington, VA: 2022

28. Guerchet M, M’belesso P, Mouanga AM, et al. Prevalence of dementia in elderly living in two cities of Central Africa: the EDAC survey. Dement Geriatr Cogn Disord. 2010;30(3):261–268. doi:10.1159/000320247

29. Ikanga J, Jean K, Medina P, Patel SS, Schwinne M, Epenge E, Gikelekele G, Tshengele N, Kavugho I, Mampunza S, Mananga L, Teunissen CE, Stringer A, Rojas JC, Chan B, Lago AL, Kramer JH, Boxer AL, Jeromin A, Gross AL, Alonso A. Preliminary reference values for Alzheimer’s disease plasma biomarkers in Congolese individuals with and without Alzheimer’s disease. medRxiv [Preprint]. 2024 Aug 7:2024.08.06.24311577. doi: 10.1101/2024.08.06.24311577. PMID: 39211852; PMCID: PMC11361236.

30. Ikanga J, Hickle S, Schwinne M, et al. Association Between Hippocampal Volume and African Neuropsychology Memory Tests in Adult Individuals with Probable Alzheimer’s Disease in Democratic Republic of Congo. J Alzheimers Dis. 2023;96(1):395–408. doi:10.3233/JAD-230206

31. Wahlund LO, Barkhof F, Fazekas F, et al. A new rating scale for age-related white matter changes applicable to MRI and CT. Stroke. 2001;32(6):1318–1322. doi:10.1161/01.str.32.6.1318

32. Claus JJ, Staekenborg SS, Holl DC, et al. Practical use of visual medial temporal lobe atrophy cut-off scores in Alzheimer’s disease: Validation in a large memory clinic population. Eur Radiol. 2017;27(8):3147–3155. doi:10.1007/s00330-016-4726-3

33. Enkirch SJ, Traschütz A, Müller A, et al. The ERICA Score: An MR Imaging-based Visual Scoring System for the Assessment of Entorhinal Cortex Atrophy in Alzheimer Disease. Radiology. 2018;288(1):226–333. doi:10.1148/radiol.2018171888

34. Kowall B, Rathmann W. HbA1c for diagnosis of type 2 diabetes. Is there an optimal cut point to assess high risk of diabetes complications, and how well does the 6.5% cutoff perform?. Diabetes Metab Syndr Obes. 2013;6:477–491. doi:10.2147/DMSO.S39093

35. Li J, Somers VK, Gao X, et al. Evaluation of Optimal Diastolic Blood Pressure Range Among Adults with Treated Systolic Blood Pressure Less Than 130 mm Hg. JAMA Netw Open. 2021;4(2):e2037554. Published 2021 Feb 1. doi:10.1001/jamanetworkopen.2020.37554

36. Whelton PK, Carey RM, Aronow WS, et al. 2017 ACC/AHA/AAPA/ABC/ACPM/AGS/APhA/ASH/ASPC/NMA/PCNA Guideline for the Prevention, Detection, Evaluation, and Management of High Blood Pressure in Adults: A Report of the American College of Cardiology/American Heart Association Task Force on Clinical Practice Guidelines [published correction appears in Hypertension. 2018 Jun;71(6):e140-e144. doi: 10.1161/HYP.0000000000000076]. Hypertension. 2018;71(6):e13–e115. doi:10.1161/HYP.0000000000000065

37. Nantsupawat N, Booncharoen A, Wisetborisut A, et al. Appropriate Total cholesterol cut-offs for detection of abnormal LDL cholesterol and non-HDL cholesterol among low cardiovascular risk population. Lipids Health Dis. 2019;18(1):28. doi:10.1186/s12944-019-0975-x

38. https://www.alz.org/media/Documents/2025-August_January-RFA_Core-Program_Final-docx.pdf

39. 2023 Alzheimer’s disease facts and figures. Alzheimers Dement. 2023;19(4):1598–1695. doi:10.1002/alz.13016

40. Nichols E, Steinmetz JD, Vollset SE, et al. Estimation of the global prevalence of dementia in 2019 and forecasted prevalence in 2050: an analysis for the Global Burden of Disease Study 2019. Lancet Public Health. 2022;7(2):e105–25. doi:10.1016/S2468-2667(21)00249-8

41. Jack CR, Therneau TM, Weigand SD, Wiste HJ, Knopman DS, Vemuri P, Lowe VJ, Mielke MM, Roberts RO, Machulda MM, Graff-Radford J. Prevalence of biologically vs clinically defined Alzheimer spectrum entities using the National Institute on Aging–Alzheimer’s Association research framework. JAMA Neurology. 2019;76(10):1174–83. doi:10.1001/jamaneurol.2019.1971

42. Young AL, Marinescu RV, Oxtoby NP, et al. Uncovering the heterogeneity and temporal complexity of neurodegenerative diseases with Subtype and Stage Inference. Nat Commun. 2018;9(1):4273. doi:10.1038/s41467-018-05892-0

43. Liss JL, Seleri Assunção S, Cummings J, et al. Practical recommendations for timely, accurate diagnosis of symptomatic Alzheimer’s disease (MCI and dementia) in primary care: a review and synthesis. J Intern Med. 2021;290(2):310–334. doi:10.1111/joim.13244

44. Giampietri L, Belli E, Beatino MF, et al. Fluid Biomarkers in Alzheimer’s Disease and Other Neurodegenerative Disorders: Toward Integrative Diagnostic Frameworks and Tailored Treatments. Diagnostics (Basel). 2022;12(4):796. doi:10.3390/diagnostics12040796

45. Hu Y, Kirmess KM, Meyer MR, et al. Assessment of a Plasma Amyloid Probability Score to Estimate Amyloid Positron Emission Tomography Findings Among Old adults With Cognitive Impairment. JAMA Netw Open. 2022;5(4):e228392–e228392. doi:10.1001/JAMANETWORKOPEN.2022.8392

46. Kirmess K, Meyer M, Holubasch M, … SKCC, 2021 undefined. The PrecivityAD^TM^ test: Accurate and reliable LC-MS/MS assays for quantifying plasma amyloid beta 40 and 42 and apolipoprotein E proteotype for the. *Elsevier*. Accessed January 19, 2023. https://www.sciencedirect.com/science/article/pii/S0009898121001601

47. West T, Kirmess KM, Meyer MR, et al. A blood-based diagnostic test incorporating plasma Aβ42/40 ratio, ApoE proteotype, and age accurately identifies brain amyloid status: findings from a multi cohort validity analysis. Mol Neurodegener. 2021;16(1). doi:10.1186/S13024-021-00451-6

48. Ikanga J, Patrick SD, Schwinne M, Patel SS, Epenge E, Gikelekele G, Tshengele N, Kavugho I, Mampunza S, Yarasheski KE, Teunissen CE, Stringer A, Levey A, Rojas JC, Chan B, Lario Lago A, Kramer JH, Boxer AL, Jeromin A, Alonso A, Spencer RJ. Sensitivity of the African neuropsychology battery memory subtests and learning slopes in discriminating APOE 4 and amyloid pathology in adult individuals in the Democratic Republic of Congo. Front Neurol. 2024 Mar 27;15:1320727. doi: 10.3389/fneur.2024.1320727. PMID: 38601333; PMCID: PMC11004441.

49. Ikanga J, Patel SS, Roberts BR, Schwinne M, Hickle S, Verberk IMW, Epenge E, Gikelekele G, Tshengele N, Kavugho I, Mampunza S, Yarasheski KE, Teunissen CE, Stringer A, Levey A, Alonso A. Association of plasma biomarkers with cognitive function in persons with dementia and cognitively healthy in the Democratic Republic of Congo. Alzheimers Dement (Amst). 2023 Nov 8;15(4):e12496. doi: 10.1002/dad2.12496. PMID: 37954546; PMCID: PMC10632676.

